# Augmentation of ChatGPT with Clinician-Informed Tools Improves Performance on Medical Calculation Tasks

**DOI:** 10.1101/2023.12.13.23299881

**Authors:** Alex J Goodell, Simon N Chu, Dara Rouholiman, Larry F Chu

## Abstract

Prior work has shown that large language models (LLMs) have the ability to answer expert-level multiple choice questions in medicine, but are limited by both their tendency to hallucinate knowledge and their inherent inadequacy in performing basic mathematical operations. Unsurprisingly, early evidence suggests that LLMs perform poorly when asked to execute common clinical calculations. Recently, it has been demonstrated that LLMs have the capability of interacting with external programs and tools, presenting a possible remedy for this limitation. In this study, we explore the ability of ChatGPT (GPT-4, November 2023) to perform medical calculations, evaluating its performance across 48 diverse clinical calculation tasks. Our findings indicate that ChatGPT is an unreliable clinical calculator, delivering inaccurate responses in one-third of trials (n=212). To address this, we developed an open-source clinical calculation API (openmedcalc.org), which we then integrated with ChatGPT. We subsequently evaluated the performance of this augmented model by comparing it against standard ChatGPT using 75 clinical vignettes in three common clinical calculation tasks: Caprini VTE Risk, Wells DVT Criteria, and MELD-Na. The augmented model demonstrated a marked improvement in accuracy over unimproved ChatGPT. Our findings suggest that integration of machine-usable, clinician-informed tools can help alleviate the reliability limitations observed in medical LLMs.

## 1 Introduction

Large language models (LLMs) such as ChatGPT and MedPaLM have demonstrated competency in the use and application of clinical knowledge, recently answering 90% of US medical licensing exam (USMLE) questions correctly [1, 2, 3]. Unfortunately, confabulated material (“hallucination”) remains a persistent problem in general and specialized domains of knowledge [4]. In addition, performing mathematical operations is challenging for large language models as they are optimized for natural language and cannot routinely perform computation with traditional methods. Calculation abilities in clinical contexts has not been thoroughly investigated, but early evidence suggests poor performance, consistent with their known shortcomings in basic math. [5].

One proposed solution to improve the calculation abilities of LLMs involves the use of semi-autonomous agent models. The concept of LLMs as autonomous agents emerged in 2022, demonstrating models’ abilities to conceive, plan, and execute tasks [6]. Recent developments have shown that they can learn to use tools, such as conducting web searches or interacting with application program interfaces (APIs) for the exchange of knowledge [7, 8]. These tool-augmented language models (TALM) have shown much superior performance in mathematics benchmarks. Instead of using their own neural network architecture to answer the question, they generate a script in a common programming language, execute that script, interpret the results, and relay the solution to the user [9]. In 2023, ChatGPT, a large language model developed by OpenAI, was enhanced with some of this functionality, including the ability to write and execute its own programs for solving complex prompts. This was known as the “code interpreter” functionality. In early November 2023, this functionality (in addition to a web-browsing tool) was integrated into the standard commercial version of ChatGPT4.

Not unlike language models, clinicians often require external tools for complex calculations. One of the most popular services, MDCalc, has between three and five million visits per month and is used by 65% of physicians in the US [10]. Its appeal lies in its focus on usability, simplicity, and safety features [11]. However, even the most intuitive applications can become burdensome if multiple calculations are required or if they require the entry of a large amount of data. This is especially true in perioperative medicine, where risk calculators such as the NSQIP or STS Risk model require 20 or more input parameters. [12, 13].

Large language models such as ChatGPT also offer a userfriendly interface through their ability to understand natural language in both written and spoken forms, and are poised become the “universal operating systems” of modern computing [14]. As these models become increasingly integrated into clinical settings and gain access to extensive patient data, they may be able to offer evidence-based calculations to clinicians [15]. It is therefore essential to understand the capabilities of LLMs to perform common clinical calculations, identify their limitations, and develop possible solutions to address these shortcomings.

In this study, we explore the performance of the mid-November 2023 version ChatGPT on 48 diverse clinical calculation tasks. We then introduce OpenMedCalc, an open-source clinical calculation API developed by the authors. This is subsequently integrated into ChatGPT and its performance is assessed against Preprint – Augmentation of ChatGPT with Clinician-Informed Tools Improves Performance on Medical Calculation Tasks 2 unimproved ChatGPT. To our knowledge, this is the first study describing the use of a tool by a language model in medicine, and the first to incorporate a clinical calculator into a language model.

## 2 Methods

### 2.1 Exploratory analysis

The initial phase of this work entailed a broad assessment of ChatGPT’s performance on a variety of calculation tasks it its unenhanced, “out-of-the-box” state, also referred to as the “base model”. The following steps were undertaken:

#### Choice of calculation tasks

To test the capabilities of ChatGPT on clinical calculation, we reviewed the 50 most popular tools offered by MDCalc and evaluated their appropriateness for the task of model assessment (Figure 1). Calculators were categorized into five types: descriptive calculators (which focus on describing a known but difficult-to-measure clinical entity, such as an anion gap or a patient’s renal function), predictive calculators (which attempt to predict the risk of some future event such as VTE), speculative (which are primarily diagnostic and focus on determining whether an entity, such as sleep apnea, currently exists in a patient), summative (which provide an overview of a disease severity or course), and therapeutic (which are meant to guide treatment in some manner). The specific calculators are shown in Table 1. Each category reflects a specific type of task that the calculator is designed to perform within a medical context. Of the 50 calculators, two were superseded by more recent calculators and were excluded. The remaining 48 calculators underwent assessment.

**Table 1:**
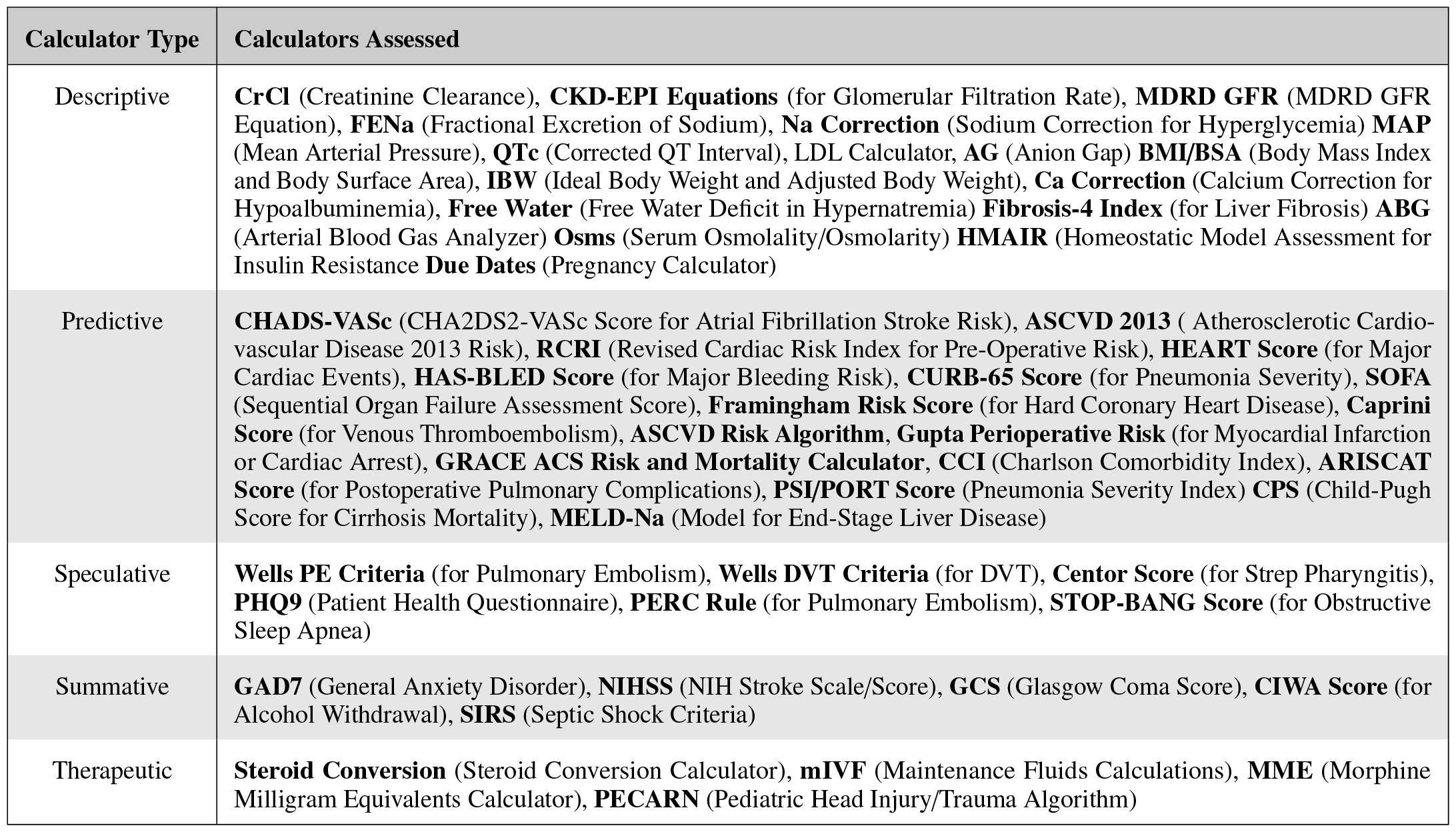
List of calculators assessed, by calculator type. Calculators were broken into five types: (1) descriptive, which attempt to estimate the value of an unmeasurable or difficult-to-measure known physical property, (2) speculative, which attempt to estimate the probability of an active disease or process. (3) summative, which combines multiple metrics to summarize the severity of an active, known disease (4) predictive, which estimates the probability or risk of a future event, and (5) therapeutic, which calculates or assists in calculation of a treatment plan.

| Calculator Type | Calculators Assessed |
| --- | --- |
| Descriptive | **CrCl** (Creatinine Clearance), **CKD-EPI Equations** (for Glomerular Filtration Rate), **MDRD GFR** (MDRD GFR Equation), **FENa** (Fractional Excretion of Sodium), **Na Correction** (Sodium Correction for Hyperglycemia) **MAP** (Mean Arterial Pressure), **QTc** (Corrected QT Interval), LDL Calculator, **AG** (Anion Gap) **BMI/BSA** (Body Mass Index and Body Surface Area), **IBW** (Ideal Body Weight and Adjusted Body Weight), **Ca Correction** (Calcium Correction for Hypoalbuminemia), **Free Water** (Free Water Deficit in Hypernatremia) **Fibrosis-4 Index** (for Liver Fibrosis) **ABG** (Arterial Blood Gas Analyzer) **Osms** (Serum Osmolality/Osmolarity) **HMAIR** (Homeostatic Model Assessment for Insulin Resistance) **Due Dates** (Pregnancy Calculator) |
| Predictive | **CHADS-VASc** (CHA2DS2-VASc Score for Atrial Fibrillation Stroke Risk), **ASCVD 2013** (Atherosclerotic Cardiovascular Disease 2013 Risk), **RCRI** (Revised Cardiac Risk Index for Pre-Operative Risk), **HEART Score** (for Major Cardiac Events), **HAS-BLED Score** (for Major Bleeding Risk), **CURB-65 Score** (for Pneumonia Severity), **SOFA** (Sequential Organ Failure Assessment Score), **Framingham Risk Score** (for Hard Coronary Heart Disease), **Caprini Score** (for Venous Thromboembolism), **ASCVD Risk Algorithm**, **Gupta Perioperative Risk** (for Myocardial Infarction or Cardiac Arrest), **GRACE ACS Risk and Mortality Calculator**, **CCI** (Charlson Comorbidity Index), **ARISCAT Score** (for Postoperative Pulmonary Complications), **PSI/PORT Score** (Pneumonia Severity Index) **CPS** (Child-Pugh Score for Cirrhosis Mortality), **MELD-Na** (Model for End-Stage Liver Disease) |
| Speculative | **Wells PE Criteria** (for Pulmonary Embolism), **Wells DVT Criteria** (for DVT), **Centor Score** (for Strep Pharyngitis), **PHQ9** (Patient Health Questionnaire), **PERC Rule** (for Pulmonary Embolism), **STOP-BANG Score** (for Obstructive Sleep Apnea) |
| Summative | **GAD7** (General Anxiety Disorder), **NIHSS** (NIH Stroke Scale/Score), **GCS** (Glasgow Coma Score), **CIWA Score** (for Alcohol Withdrawal), **SIRS** (Septic Shock Criteria) |
| Therapeutic | **Steroid Conversion** (Steroid Conversion Calculator), **mIVF** (Maintenance Fluids Calculations), **MME** (Morphine Milligram Equivalents Calculator), **PECARN** (Pediatric Head Injury/Trauma Algorithm) |

**Figure 1:**
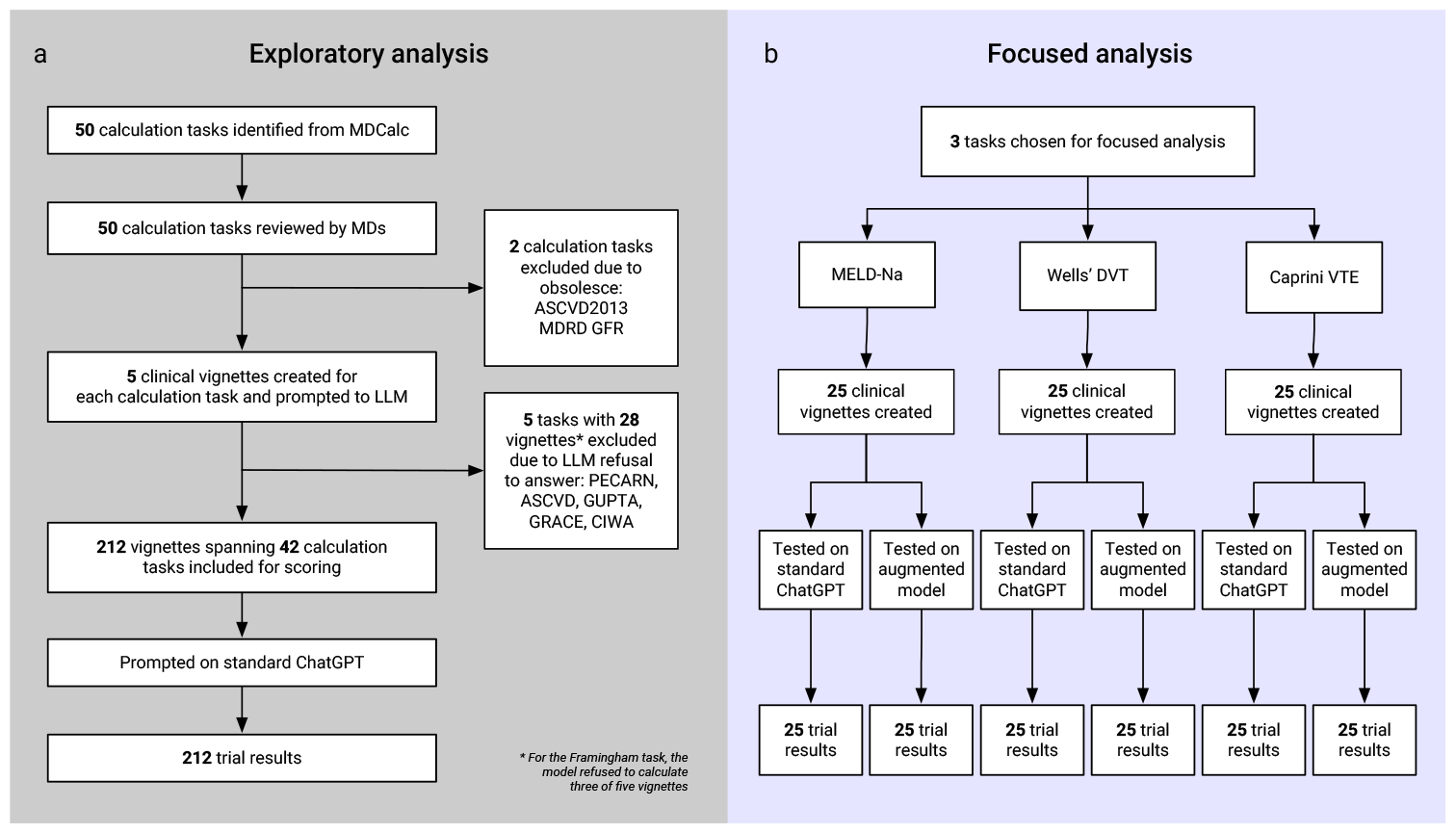
Overview of study’s approach to assessment of clinical calculation. The schematic delineates the two-phase process. **(a)** Exploratory analysis of 50 calculation tasks identified from MDCalc. Tasks were reviewed and vetted for suitability, resulting in 212 vignettes spanning 42 tasks after exclusions due to obsolescence or LLM’s inability to provide answers. **(b)** In the focused analysis, in-depth testing of the LLM’s performance on the Caprini VTE, Wells DVT, and MELD-Na tasks was performed with both the base and augmented models, across a set of 25 clinical vignettes per task.

#### Vignettes

After review of the calculator, a convenience sample of five clinical vignettes were written by one of the two physician authors. The vignettes were meant to relay all necessary information to perform the calculation with minimal extraneous information, and contain a request for a calculation that was unambiguous. To minimize question-answering avoidance (i.e. “as an AI model I am not…”), prompts were written from the perspective of a clinician asking for assistance, using commonplace medical jargon. In cases where multiple common formulas existed for a calculation and no clear gold standard was established, we specified a preferred method (i.e. “calculate the QTc using the Bazett formula”). Data were presented in an objective fashion as much as possible. Where relevant, continuous values were reported within a range that one would reasonably see in clinical practice; units were omitted in most prompts, as is common in clinical communication. For criteria-based calculators, the vignettes provided the underlying data as opposed to whether criteria were met. For example, a vignette for CHA_2_DS_2_VASc might report a “history of TIA,” not “positive cerebrovascular disease.” The intent was to convey information in manner that was clear from a clinical perspective, but indirect, simulating how a clinician might use the tool.

#### Presentation and scoring

To evaluate the recently-added dynamic programming abilities in ChatGPT, prompts were presented to the web interface of the commercial version of Chat-GPT by one of two physician assessors. Each vignette was started in its own conversation to eliminate contamination from prior prompts. If the LLM did not provide a definitive answer but showed progress towards an answer, the user encouraged the conversation with a prompt such as “Please continue.” To score the performance of the model, assessors compared Chat-GPT’s answers with the answer provided by MDCalc with input data from the vignette. For criteria-based models, answers were counted as correct if the reported scores matched. For models with continuous variable outputs, rounding errors were permitted if the calculator did not specify how to round. Some of the calculators involved calculations without a gold standard, such as drug conversions. Responses on these models were considered correct if the underlying math was executed appropriately and the dose given was reasonable. If a model was unable or refused to answer a question even after encouragement with two prompts, a score of “unable to calculate” was given. The conversation was saved to a database in plain text. When available, a URL for the conversation was saved. These conversations have been provided in an accessible format online [16].

#### Classification of errors

After initial trials were complete, we inspected the correct and incorrect answers. From these, we formulated a framework for the chain of reasoning required to provide appropriate answers, as well as the various possible missteps within this chain (Figure 2). When the model offered incorrect solutions, raw output was inspected for these errors in reasoning; follow-up questions were not asked. Errors were categorized into one (or more) of categories listed below. After classifying all errors, Pearson’s correlations were calculated to determine the relationship between calculator type and error class.

**Figure 2:**
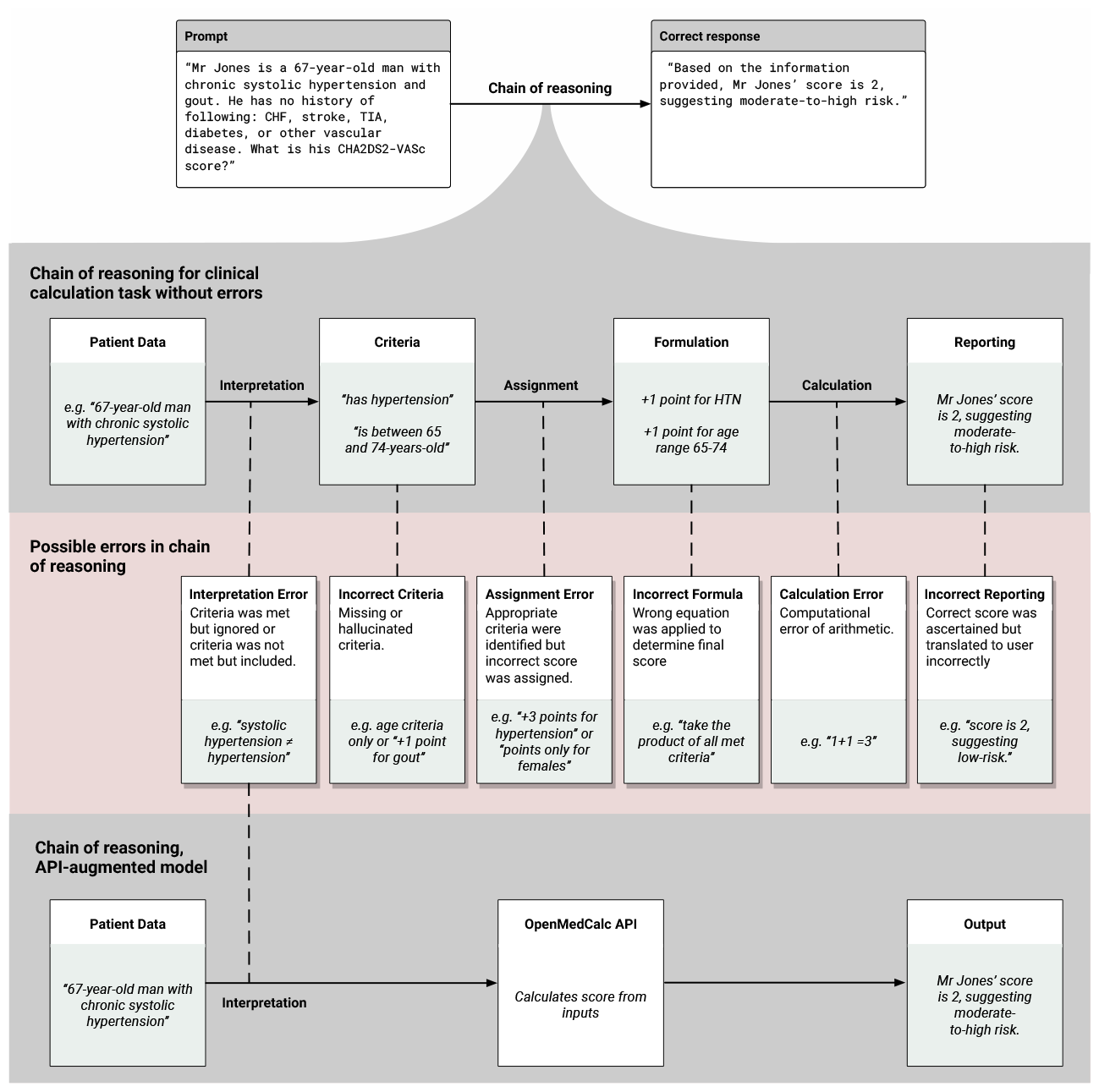
Chain of reasoning and error classification framework with example CHA_2_**DS**_2_VASc prompt. This diagram provides a detailed overview of the approach used to classify errors encountered in the clinical calculation tasks performed by LLMs. It shows an example prompt with the user asking to calculate a CHA_2_DS_2_VASc for a patient, and the correct response from the model. The second row delineates the necessary reasoning steps required to accurately complete a clinical calculation manually, shown as a row of connected processes with examples of correct conclusions from the CHA_2_DS_2_VASc example prompt. The third row illustrates the possible errors in this chain of thought, shown as call-out boxes with example errors, linked to the respective step in the chain. The final row shows the same chain of reasoning as the first, but with the inclusion of a calculation API, thereby bypassing many of the steps involved in calculation and eliminating the chance of error. Note only interpretation errors are linked to the API-augmented approach.

- **Interpretation Error** Inadequate understanding or misinterpretation of the medical information presented in the question leads to 1) ignored criteria that was met, or 2) inclusion of a criteria that was not met.
- **Incorrect Criteria** Criteria are missing or there is a hallucination of non-existent criteria.
- **Assignment Error** Improper application of correctly-identified criteria. Appropriate criteria were selected but an incorrect score is assigned.
- **Incorrect Formula** An incorrect equation is chosen to represent the scoring mechanism of the calculation task.
- **Calculation Error** The correct formula is chosen, such as taking the sum of all subscores, but the actual mathematical computation carried out was incorrect.
- **Incorrect Reporting** The correct score is calculated, but some component of reporting that score to the user is inaccurate.

### 2.2 Development and Evaluation of an Open Medical Calculator

After completion of the exploratory phase, we developed a traditional computational calculator and integrated it within a custom version of ChatGPT using the following steps:

#### Calculator development

A basic application program interface (API) was created using FastAPI [17] and installed on a remote server [18]. Three calculation tasks from the exploratory analysis were chosen as representative of ChatGPT’s weaknesses. For each of these tasks, a algorithm was written and implemented within the API (Python v3.11, Portland, OR). The algorithms included upper and lower bounds for inputs as well as outputs. The API and its documentation conformed to OpenAPI specification, an open source effort to standardize machine-readable communication [19]. The codebase for the algorithms and API are accessible online [20].

#### Integration with ChatGPT

Using commercially-available ChatGPT plus account, a “custom GPT” was created using the web interface on OpenAI’s platform (Figure 4). A system prompt was added to further instruct the chatbot: “You are a helpful medical calculation bot. Rely on the provided medical calculation APIs to answer the user’s questions. If data is missing, do not make assumptions but instead ask clarifying question.” This custom bot is available online to users with ChatGPT Plus subscriptions [21].

#### Focused prompting

After basic testing and debugging of the API, we compared the base model with the API-augmented approach. To do so, a program was written to generate 25 case vignettes with randomly-selected relevant patient attributes for the three different calculators, forming a total of 75 unique case vignettes. The primary outcome of interest was percentage correct on each calculation task. The sample size of 25 was chosen arbitrarily, based on available resources. Each vignette was structured identically, containing both the primary relevant data and supplemental information necessary to establish a clinical context. Example of these vignettes are included for reference in Table 2. Each of the 75 vignettes was then used to prompt the base model followed by the API-augmented model, resulting in a total of 150 trials. The methods used in prompting either the base model or augmented model were identical to those described in Section 2.1. Vignettes and model responses are accessible online [16].

**Table 2:**
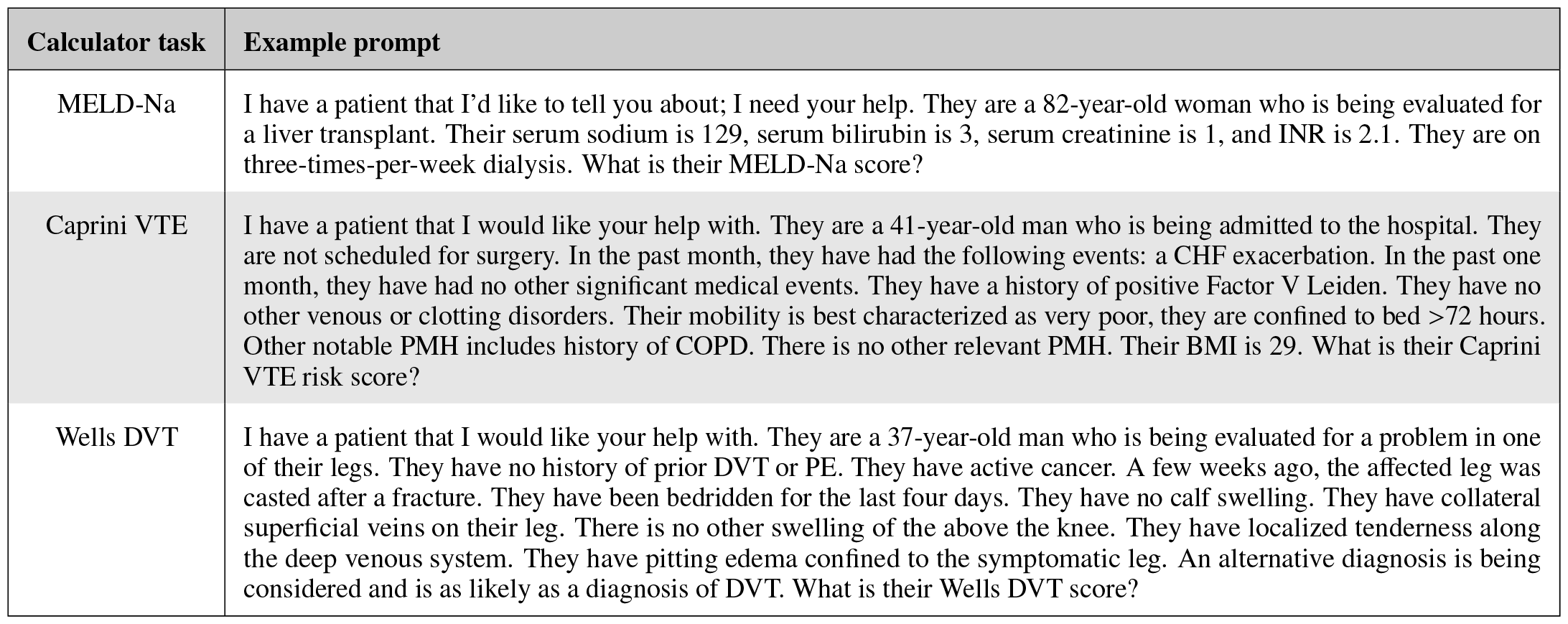
Example vignettes. Three example vignettes used for prompting during the focused analysis.

| Calculator task | Example prompt |
| --- | --- |
| MELD-Na | I have a patient that I'd like to tell you about; I need your help. They are a 82-year-old woman who is being evaluated for a liver transplant. Their serum sodium is 129, serum bilirubin is 3, serum creatinine is 1, and INR is 2.1. They are on three-times-per-week dialysis. What is their MELD-Na score? |
| Caprini VTE | I have a patient that I would like your help with. They are a 41-year-old man who is being admitted to the hospital. They are not scheduled for surgery. In the past month, they have had the following events: a CHF exacerbation. In the past one month, they have had no other significant medical events. They have a history of positive Factor V Leiden. They have no other venous or clotting disorders. Their mobility is best characterized as very poor, they are confined to bed >72 hours. Other notable PMH includes history of COPD. There is no other relevant PMH. Their BMI is 29. What is their Caprini VTE risk score? |
| Wells DVT | I have a patient that I would like your help with. They are a 37-year-old man who is being evaluated for a problem in one of their legs. They have no history of prior DVT or PE. They have active cancer. A few weeks ago, the affected leg was casted after a fracture. They have been bedridden for the last four days. They have no calf swelling. They have collateral superficial veins on their leg. There is no other swelling of the above the knee. They have localized tenderness along the deep venous system. They have pitting edema confined to the symptomatic leg. An alternative diagnosis is being considered and is as likely as a diagnosis of DVT. What is their Wells DVT score? |

#### Focused analysis

After completion of the 150 trials of the selected calculators, the models’ responses were analyzed in a method similar to that described above, including classification of errors for responses that were scored as incorrect. To compare the performance of the base model to the augmented model, a Chi-squared test was utilized. All analysis was performed on Jupyter notebooks using Python 3.11.

## 3 Results

### 3.1 Findings from exploratory analysis

Of the 48 calculation tasks tested, a total of six tasks generated atypical answers. ChatGPT was unable to give any substantial answer to four tasks: the GRACE, GUPTA, PECARN, and ASCVD (Figure 1). For these prompts, the LLM browsed the internet to find human-usable medical calculators such as MDCalc and offered links to the user as suggestions. For the calculation of CIWA scores, which involve subjective grading of physical exam findings such as perceived anxiety or tremor, the LLM would only provide ranges. Thus, these five tasks were graded “unable to calculate.” For the Framingham Score, three trials resulted in ChatGPT indicating its inability to calculate the score, instead providing links to relevant materials. In two trials, it attempted to provide an answers by hallucinating a calculation scheme, one of which resulted in a myocardial infarction risk estimate near 1% and a second with a risk over 100% for similar patients [16]. The first three attempts were coded as “unable to calculate,” while the latter two attempts were scored as incorrect.

Out of the remaining 42 calculators, ChatGPT successfully provided calculations in five of five trials, resulting in a total of 212 scorable answers. Of these, 140 were classified as correct, representing 66% of all trials. There was considerable variation in performance across calculation tasks; in six tasks, none of the provided answers were correct, while in 17 tasks, all provided answers were correct (Figure 3). There was a notable difference based on calculator role: for tasks classified as “predictive”, the model was able to accurately answer only 39% of trials, while calculation tasks classified as “descriptive” were answered correctly in 89% of trials (Figure 3).

**Figure 3:**
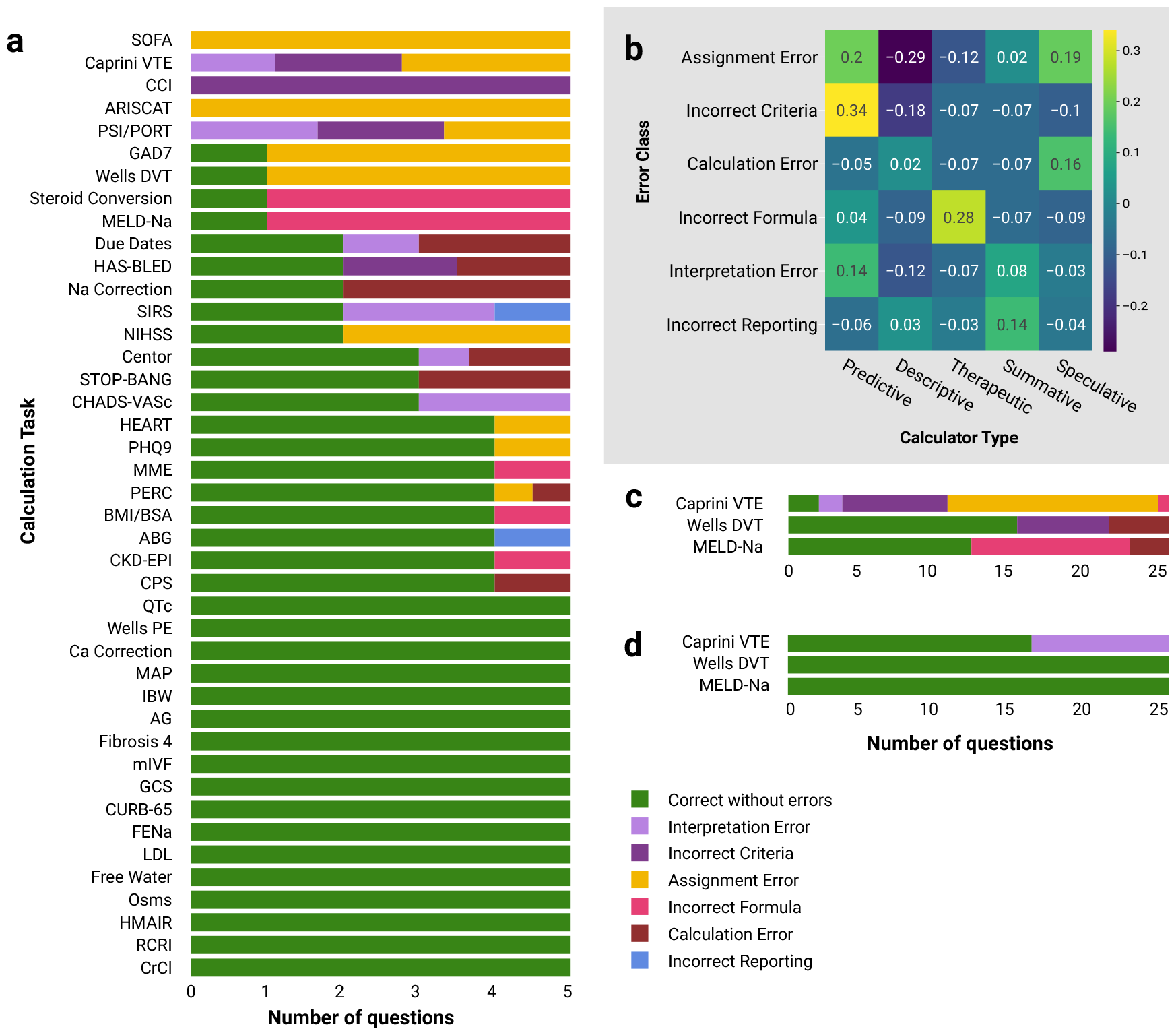
Detailed breakdown of ChatGPT’s performance on calculation tasks. **(a)** Vertical histogram of number of questions answered correctly (green), sorted by calculation task in exploratory analysis. Number of correct answers shown in green, colored bars other than green signify errors, with the class specified by color. **(b)** Heatmap of Pearson correlation coefficients between calculator type and error classes identified in exploratory analysis. This heatmap visualizes the strength and direction of relationships between the different roles of calculators—predictive, descriptive, therapeutic, summative, and speculative against the classes of errors made by the base model. Blue shades indicate stronger negative correlations, while green shades indicate stronger positive correlations. Notably, descriptive calculation tasks correlate with fewer assignment errors, while predictive tasks show a higher propensity for both assignment and incorrect formula errors, reflecting the complexity of the task and the models lack of familiarity. **(c)** Vertical histogram, similar to that of subplot a, showing correct answers for the three tasks evaluated in the focused analysis using the base model as well as **(d)** using the augmented model.

**Figure 4:**
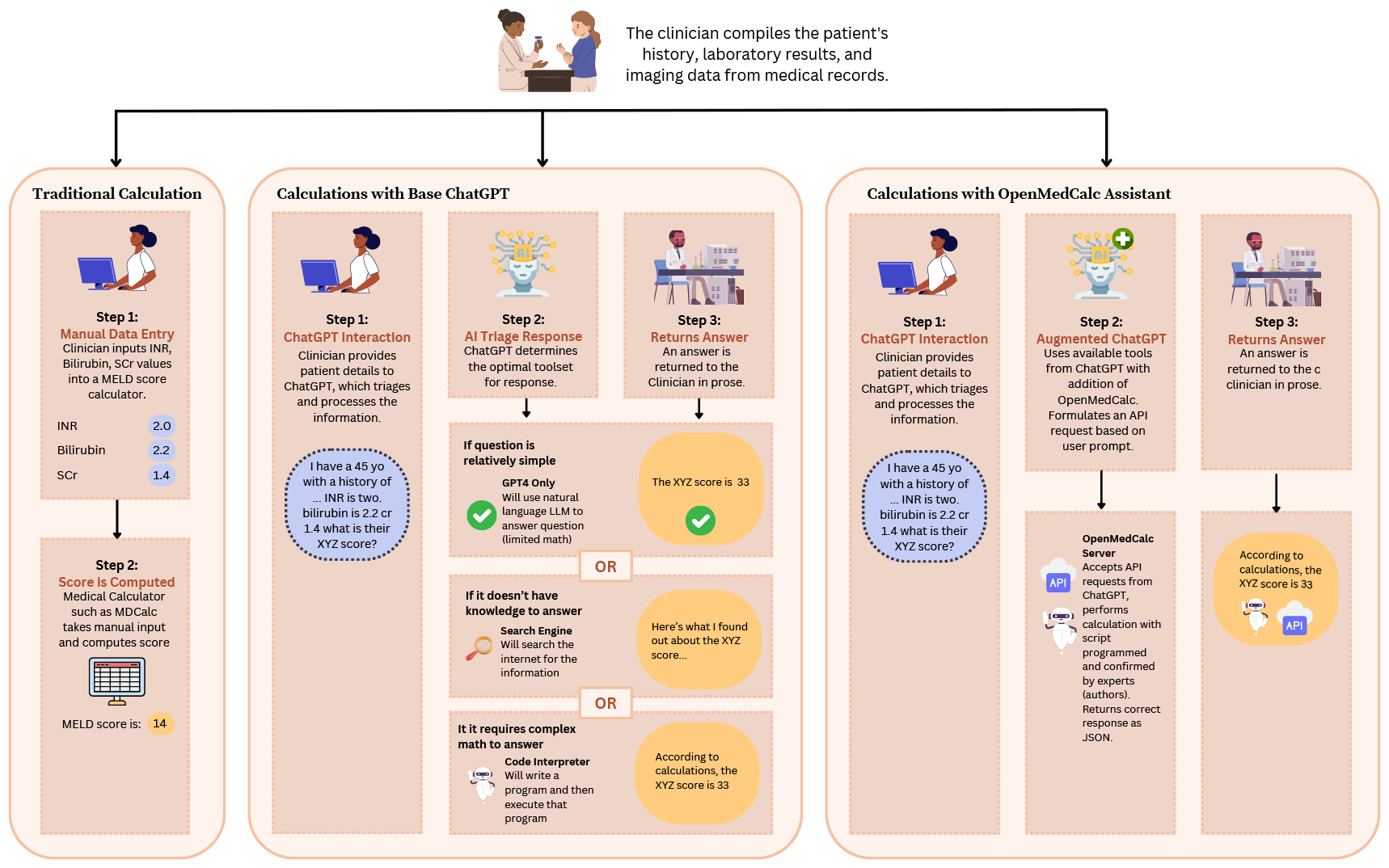
Overview of workflow for clinical calculation across three methods of calculation. This schematic delineates the sequential flow of data and decision-making processes across three distinct clinical calculation scenarios: traditional manual calculation, automated calculation with the base model (standard ChatGPT), and enhanced calculation using the API-augmented model (OpenMedCalc assistant). The traditional approach involves manual data entry and score computation through a web-based medical calculator. The base ChatGPT method triages the prompt to decide the optimal tools in developing a response, with contingencies for simple queries, knowledge gaps, and complex math requirements. The OpenMedCalc-augmented pathway showcases an advanced integration where ChatGPT leverages a specialized API to perform accurate clinical calculations, reflecting a blend of AI efficiency with expert-validated computational precision.

### 3.2 Error classification from exploratory analysis

During the exploratory analysis, there were a wide array of errors made by ChatGPT in its attempts to answer clinical calculation tasks; a total of 84 errors were identified across 72 questions. The most common errors were *assignment errors* (31 of 82 errors, 38% of errors), where the model correctly identified the criteria but assigned the wrong score for that criteria. This was more common in longer calculation tasks which had more criteria, making it difficult for the model to “recall” the correct points for each criterion. For example, in the ARISTCAT task, the model identified that preoperative SpO_2_ was a part of the criteria, but was inconsistent on how many points an abnormal value contributed to a final score.

The second most common error class was *incorrect criteria*, where the model either hallucinated new criteria or omitted relevant criteria; this comprised 13 of 82 identified errors (16%). Without a clear understanding of criteria, the model would hallucinate variations of criteria from similar calculators, many of which would appear reasonable to an unfamiliar user. A pointed example comes from the model’s attempt to assess an individual’s Caprini VTE score, where it informed the user that a young overweight patient did not score any points for being overweight because “‘*BMI is only counted as a risk factor in patients aged over 40*.”

The third and fourth most common errors classes involved the application of math within the model. When presented with a calculation task, the base model would either (1) attempt to calculate using the language model alone, or (2) use the “‘code interpreter” tool. In the former, the model displayed difficulties performing simple calculations and/or counting. In one HAS-BLED prompt, it was unable to calculate 1+1. These were classified as *calculation errors* and comprised 13 of 82 errors (16%). The later, code interpreter functionality was reserved more complex math. In general, the model would describe the formulas in plain text or forumla notation. Subsequently, it would pause and proceed to write a program in Python to calculate the results. The delivered result was occasionally incorrect even in instances where the model presented a correct version of the formula prior to writing the script. These were classified as having an *incorrect formula*, and represented 12 (15%) of the identified errors. Interpretation errors (11 of 82, 14%) were also common. In particular, the base ChatGPT model occasionally struggled with interpreting numerical ranges, for example, reporting that a 64-year-old man would be categorized as between “65-74” in calculating a CHA_2_DS_2_VASc score or that a partial pressure of oxygen of 66 was “less than 60.”

After identification of the errors, the classes of errors were compared to the calculation method and role (Figure 3). Two patterns emerged from analysis of the errors. Firstly, the base model was less likely to make an assignment error in the “descriptive” models (such as estimating creatinine clearance or correcting hypercalcemia for albumin levels). This was likely due to the simple assignments required by these calculators; there is not a complex scoring system derived from a linear regression. In contrast, the model was more likely to make assignment errors or use the incorrect formula with “predictive” models such as the SOFA tool. This reflects the higher level of complexity in the criteria of these models, as well as the base models’ lack of familiarity with some of these calculators.

### 3.3 Focused analysis in standard ChatGPT

Our exploratory analysis revealed that ChatGPT was inaccurate in one-third of clinical calculations due to errors in knowledge and logic. To address this, we developed an API to assist Chat-GPT in routine clinical calculations. To test this solution, we further characterized three clinical tasks where ChatGPT’s performance was weakest: the Caprini VTE risk score (0 out of 5 correct), the Wells’ score for DVT (1 out of 5 correct), and the MELD-Na score (1 out of 5 correct). Given the small sample sizes of our exploratory analysis, we expanded our trials to 25 clinical vignettes, hoping to garner a more in-depth understanding of the base model’s capabilities and weaknesses on these tasks.

Performance of the base model on the three tasks was better than measured in the exploratory analysis, with a total of 29 correct answers from 75 prompts. The base ChatGPT4 model correctly answered 2 of the 25 trials (8%) for the Caprini VTE score, 12 of 25 trials (48%) for the MELD-Na score, and 15 of 25 trials (60%) for the Wells DVT score (Figure 3).

Similarly to the exploratory analysis, the most common error classes identified were assignment error (18, 33%) and incorrect criteria (15, 27%). These were most pronounced in the Caprini VTE risk assessment, where the model repeatedly hallucinated criteria and their assigned scores. For example, it reported various scoring mechanisms for the overweight criteria, using different scores at cutoffs of BMI of 25, 30 or 35 kg/cm2; the actual criteria has only one cutoff at 25 ([22]). The model also struggled recalling the scoring system for the clotting risk factors and hallucinated different criteria. For example, the model added one point for an elevated homocysteine, reporting that it was a *“possible indicator for thrombophilia, although not explicitly listed in the Caprini score*.*”* In reality, an elevated serum homocysteine is explicitly assigned a score of three points in the Caprini score ([22]).

The third most common error was an incorrect formula, comprising 13 of 55 errors (24%). These were primarily found in the MELD-Na trials, where the model made a variety of errors. A common error was the inappropriate combination of formula terms from previous versions of the MELD criteria, or inappropriate/omitted cutoff values for laboratory inputs. Although not formally assessed, it was notable that the model exhibited more confidence in the MELD-Na trials, rarely offering caveats that it may not be qualified, but rather relaying convincing descriptions of its methods (Figure 5).

**Figure 5:**
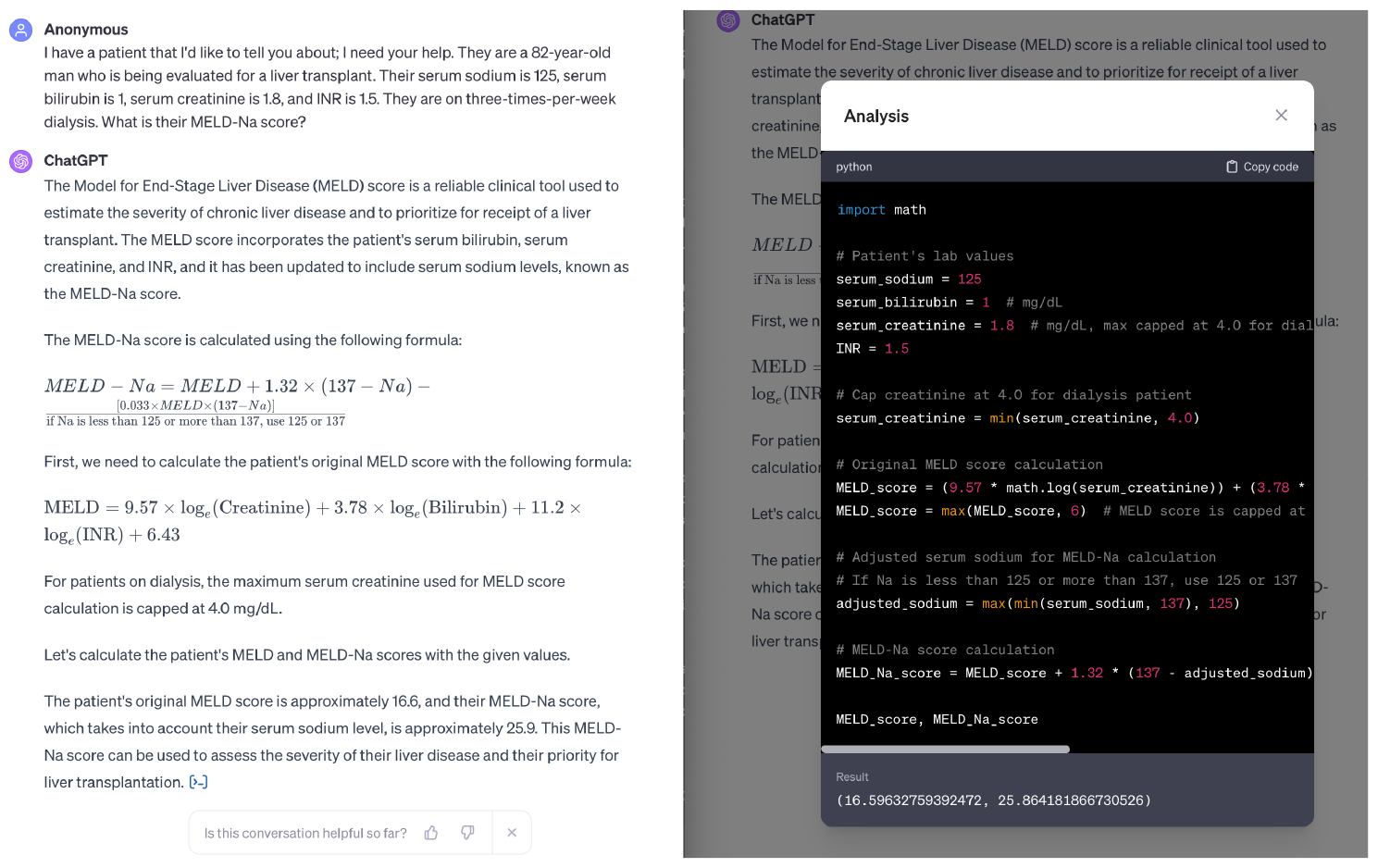
Illustration of standard ChatGPT’s response to MELD-Na prompt. The left panel outlines the inquiry about a patient’s clinical details as well as ChatGPT’s initial response, where it outputs plain text with mathematical formulas describing its plan for calculation. The right panel displays the Python code written and executed by ChatGPT to perform the calculation. Note that the creatinine adjustment method is incorrect (the creatinine should be set to 4.0 for those on dialysis, not capped at 4.0).

### 3.4 Focused analysis in API-augmented ChatGPT

As part of our focused analysis of the three chosen tasks, we built a calculator API and integrated it into an augmented version of ChatGPT. We then repeated the trials using the same vignettes. Performance was markedly improved with the addition of the API. The base model was able to answer 29 (39%) of trials correctly, while the augmented model was able to answer 66 (88%) of trials correctly (Table 3). The augmented model was able to answer all trials correctly for both MELD-Na and the Wells’ DVT calculations, eliminating hallucination in these tasks. The augmented model displayed improvement on the Caprini VTE task, correctly answering 16 of 25 (64%) trials compared to 2 of 25 (8%) in the base model (Figure 3). Each of these was statistically significant (p<0.01, Table 3).

**Table 3:** Performance of API-augmented model with base unimproved model in three calculation tasks. This includes results for the three calculation tasks used to evaluate the two models during the focued analysis, Caprini VTE, Wells’ DVT, and MELD-Na. The table shows improved performance in all tasks with a p-value of less than 0.01.

| Calculator | Model | # Correct (%) | # Questions | Notes |
| --- | --- | --- | --- | --- |
| Caprini VTE | Base model | 2 (8) | 25 | chi-squared=17.0, p-value<0.00005 |
|  | API-augmented model | 16 (64) | 25 |  |
| MELD-Na | Base model | 12 (48) | 25 | chi-squared=17.6, p-value<0.00005 |
|  | API-augmented model | 25 (100) | 25 |  |
| Wells DVT | Base model | 15 (60) | 25 | chi-squared=12.5, p-value<0.0005 |
|  | API-augmented model | 25 (100) | 25 |  |
| Total | Base model | 29 (39) | 75 | chi-squared=39.3, p-value<0.000001 |
|  | API-augmented model | 66 (88) | 75 |  |

Upon review of the nine errors found in 75 trials, all were errors of interpretation. In six, “total hip arthoplasty” was characterized as “major surgery” instead of “major lower extremity surgery.” The remaining three errors concerned interpretation of mobility, classification of major versus minor surgery, and misclassification of a radial fracture as a lower extremity fracture. Notably, there were no errors in the other classes, consistent with our framework (Figure 2).

## 4 Discussion

In this work, we demonstrate that despite access to web search and code execution tools, ChatGPT is an unreliable calculator, providing correct answers in only two-thirds of trials across a diverse group of tasks. Yet, when given access to a task-specific tool, performance showed marked improvement, increasing accuracy to 100% and eliminating hallucination in two of the three tasks assessed, while significantly improving performance in the third.

Tool-augmented language models (TALM) have been explored in fields outside of medicine, revealing that even smaller language models, when supplemented with specialized tools, can yield results comparable to those of foundational models [23]. Yet, this approach has not been explored in a medical context Preprint – Augmentation of ChatGPT with Clinician-Informed Tools Improves Performance on Medical Calculation Tasks 9 until now. Given the notable improvement observed with API integration, we propose that tool-using LLMs will be essential to many applications of machine learning within medicine and are poised to revolutionize care. Tasks like routine clinical calculations may be fully automated within the foreseeable future, as LLMs extract and analyze the relevant text data from the electronic medical record (EMR) and communicate with an API to perform these tasks. Fields such as preoperative risk assessment have the potential to be transformed as LLMs remove burdensome data entry tasks from clinicians and provide automatic, personalized risk analysis to patients.

Given the number and variety of errors encountered here, there is still significant improvement needed before this promise is to be realized. To better understand these errors, we introduced a novel classification schema based on the errors’ position within a chain of logical steps. With utilization of the calculation API, we were able to eliminate errors in five of the six error classes. Yet, the model was still susceptible to misinterpretation of the prompt. Thus, our framework may assist in the differentiation of errors susceptible to the use of an external tool, and those errors which may be intractable (Figure 2). Our framework does not, however, address the underlying cause of the errors, such as inadequate reasoning or lack of knowledge. These might be attributable to the model’s training methods, neural network structure, or reinforcement techniques, for example. To understand these causes, a different framework for error classification would be needed, as well as data from the intermediate layers of the neural network.

In addition to better characterization of the root cause of models’ errors, large-scale implementation will require large-scale evaluation. With recent work showing ChatGPT’s ability to answer 90% of USMLE-style questions correctly, the utility of multiple-choice questions as benchmarks for medical LLM evaluation may be diminishing [1]. More diverse and holistic benchmarks, such as those produced by the MedAlign and Med-HALT projects, are needed [24, 25]. Given the poor performance of unimproved ChatGPT on the tasks in this study, clinical calculation tasks may warrant inclusion in future benchmarks. Thus, we have released our vignettes and scoring metrics for this purpose [16].

There were several notable limitations to this study. As a natural language model, LLMs are unsurprisingly sensitive to variations in question phrasing. Though we established general guidelines for our exploratory analysis, there was variation in the wording of prompts. In the focused analysis, question stems were standardized to reduce this variability. These differences in phrasing likely had unpredictable and difficult-to-characterize impacts on our results. However, since our aim was to simulate how clinicians might interact with a LLM “in the wild,” under real-world settings, this variability may be appropriate. One important subset of phrasing is prompt engineering, where different key terms or styles of questioning are employed to improve LLM output. For example, prompting LLMs to articulate their step-by-step reasoning process, known as chain-of-thought (CoT) prompting, has been shown to significantly enhance performance [26, 1]. It is conceivable that prompt engineering would have improved performance of either model, as well as our ability to pinpoint the model’s reasoning [27, 28]. One technical limitation was our reliance on the web version of ChatGPT instead of the more programmer-friendly API. This was chosen to more accurately capture the real-world computational environment, as the exact model configuration parameters used in the web-based Chat-GPT are not public information. However, because the web version does not report its randomization seed, reproduction of our results is infeasible, a common challenge in AI research [29].

This work also presents many opportunities for future research. In our study, we excluded tasks that ChatGPT refused to attempt. It is possible with continued prompts or experimentation with prompt engineering, the model may have attempted these tasks. Exploring when a model refuses to answer a question outside its estimated abilities (termed “out-of-distribution problems”) is essential to LLM safety in the medical context and deserves continued exploration [30, 31]. In addition, though we classified errors in a novel framework, we did not measure the magnitude of the error in all tasks. Future studies could evaluate, for example, what proportion of errors would have lead to a change in medical management of the patient in question. Additionally, exploration of clinical calculation by multi-modal models, which posses the capability to interpret and analyze images, or multi-agent models, which simulate medical teams, may also provide insight into a different set of challenges [32, 33]. Lastly, as OpenMedCalc is an open-source tool, it can be used for studying any of the thousands of medical calculators that have been publicly described. Contributions from other clinicians, scientists, and engineers are welcome [18].

In summary, this work introduces a novel approach to improving the output of language models through the use of a clinician-informed, task-specific tool. Ultimately, LLMs stand at the cusp of revolutionizing medical practice, promising a more efficient, equitable, and patient-centered approach. In this evolving landscape, it is imperative for models to exhibit high levels of robustness and reliability. Achieving this will likely necessitate an ensemble of models and tools to effectively meet these demands, ensuring that LLMs can be integrated safely into the healthcare domain.

## Data Availability

All data produced are available online at https://github.com/stanfordaimlab/llm-as-clinical-calculator

## 5 Author contributions

AG conceived, designed, and implemented the OpenMedCalc API system, designed the methodology of the study, performed data analysis, and wrote first draft of manuscript. AG and SC conceptualized study, collected data, reviewed LLM responses for error classification, and contributed to manuscript. DR provided critical review and was a contributor to the manuscript. LC provided critical review and was a contributor to the manuscript. All authors read and approved the final manuscript.

## 6 Acknowledgements

The authors thank Graham Walker of MDCalc for discussion of concepts presented in paper.

